# Hypothesis on the neurophysiological architecture of postural control: validation in a model of Cerebral Palsy using frequency analysis

**DOI:** 10.64898/2025.12.29.25343135

**Authors:** E.V. Strelnikova, L.B. Oknina, A.A. Slezkin, A.O. Kantserova, M.E. Myachina, A.V. Kirichenko, M.V. Shtern

## Abstract

**Introduction:** Traditional clinical motor function assessment scales, despite their importance, often fail to identify the underlying neurophysiological mechanisms of postural control disorders. In this regard, stabilography, as an objective quantitative method, acquires particular diagnostic value.

The aim of this study was to identify and compare frequency markers of postural control disorders in adults with cerebral palsy (CP) and healthy subjects using stabilographic signal power analysis in narrow frequency ranges.

**Methods:** Stabilograms were recorded while performing a visual feedback task and its combination with additional cognitive loads in two groups: adults with CP (n=8) and a control group (n=8). For the analysis, the stabilographic signal power was calculated in ten narrow frequency ranges (0.05–12.0 Hz).

**Results:** Based on the analysis of the stabilographic signal power, individual postural control profiles were identified, defined by three key patterns: «hyperactivation», «exhaustion», and «optimization». The obtained data were interpreted within the framework of N.A. Bernstein’s level theory of movement construction, where the identified patterns reflect an imbalance or synergy of various postural regulation circuits—from subspinal to corticocerebellar.

**Conclusions:** The proposed method for analyzing stabilogram power enables the identification of individual neurophysiological profiles of postural control disorders. The identified «optimization» marker indicates preserved neuroplastic potential. The results of the work open the way to a well-founded personalized rehabilitation, the strategy of which consists of transforming a pathological pattern («hyperactivation», «exhaustion») into an optimal one («normalization») through targeted modulating effects on specific levels of movement construction.

## 1. Introduction

Cerebral palsy (CP) is a heterogeneous group of non-progressive motor and postural developmental disorders resulting from brain damage early in ontogenesis [1; 2]. A key factor limiting patients’ motor activity and social integration is a deficit in postural control—the ability to maintain postural stability and adequately respond to external stimuli [3; 4]. Traditional clinical assessment scales (GMFM, FMS) [5], despite their value for functional diagnostics, do not allow for the uncovering of the neurophysiological architecture of the disorders and the compensatory strategies that develop in response to the primary defect.

In this regard, objective quantitative methods, such as stabilography, which records oscillations of the center of pressure (COP) [6], are of particular importance. Spectral analysis of the stabilographic signal is considered a reliable tool for assessing postural control [7], as it allows for the decomposition of overall postural variability into frequency components traditionally associated with the activity of various sensory systems. According to the literature, the following frequency ranges correspond to the main sensory modalities: visual system: 0.02–0.2 Hz, vestibular system: 0.1–0.2 Hz, cerebellar regulation: 0.5–2.0 Hz, proprioceptive system: 1.0–5.0 Hz [8–14].

However, it should be noted that the frequency boundaries attributed to each system vary significantly across studies, and their interpretation is often not directly based on fundamental theories of motor control organization.

In this regard, objective quantitative methods, such as stabilography, which records oscillations of the center of pressure (COP) [6], are of particular importance. Spectral analysis of the stabilographic signal, in particular, is considered a reliable tool for assessing balance [7], allowing for the identification of frequency components traditionally associated with the activity of various sensory systems. Literary data indicate the following correspondences between sensory systems and frequency ranges: vision—frequencies of 0.02-0.2 Hz, vestibular system—0.1-0.2 Hz, cerebellar regulation—0.5-2.0 Hz, proprioceptive system—1.0-5.0 Hz [8–14]. It should be noted that the frequency corridor associated with a specific sensory system by various authors is quite broad, and the interpretation of frequency characteristics often does not have a strong connection with fundamental theories of motor control organization.

This work proposes a fundamentally new interpretative framework based on the synthesis of modern methods of frequency analysis of the stabilogram with the concepts of the classical Russian physiological school. Evaluation of the research results allowed us to put forward a hypothesis examining the frequency patterns of the stabilogram through the prism of movement construction levels according to N.A. Bernstein [15] and the theory of functional systems by P.K. Anokhin [16; 17]. According to this hypothesis, a disruption of a certain level of motor control (according to Bernstein) is the primary cause of postural balance control defects in cerebral palsy. In response, a compensatory functional system (according to Anokhin) is formed, aimed at achieving a beneficial result — stability. We propose that the frequency characteristics of COP oscillations are objective indicators of the properties of this reorganized system, quantitatively reflecting the dominant levels of regulation and the degree of their synergy or conflict, as well as the effectiveness and «cost» of compensation.

### 2. Methods

### 2.1. Subject Characteristics

The study included 16 subjects, divided into two age-matched groups with different levels of cerebral palsy. The characteristics of both groups are presented in Tables 1 and 2.

**Table 1.** Characteristics of healthy subjects and screening test results.

| № | Age range | Gend. | Edu. | Physical activity level | MoCA | Romberg test * |  |  | Leading hand | Leading leg |
| --- | --- | --- | --- | --- | --- | --- | --- | --- | --- | --- |
|  |  |  |  |  |  | Eyes Open | Eyes Closed | Tandem |  |  |
| H_01 | 21-25 | F | partial he | average | 29 | good | satisfact. | unsatisfat. | R | R |
| H_02 | 21-25 | F | partial he | low | 30 | good | satisfact. | unsatisfat. | R | R |
| H_03 | 21-25 | F | partial he | average | 30 | good | satisfact. | unsatisfat. | R | R |
| H_04 | 21-25 | M | partial he | low | 30 | good | unsatisfat. | unsatisfat. | R | L |
| H_05 | 21-25 | M | partial he | average | 30 | good | good | satisfact. | R | L |
| H_06 | 21-25 | F | he |  | 29 | good | satisfact. | unsatisfat. | R | R |
| H_07 | 21-25 | M | partial he | average | 30 | good | satisfact. | unsatisfat. | R | R |
| H_08 | 26-30 | F | he | high | 30 | good | satisfact. | unsatisfat. | R | R |

**Note:**
Gend. — gender
F — woman
M — man
Edu. — education
partial he — partial higher education
he — higher education
R — right
L — left
Romberg test \*. Position: The subject stands with his feet together and his arms extended in front of him at shoulder level.

**Table 2.** Characteristics and results of screening tests of study participants with cerebral palsy.

| Code | Age range | Gend. | Type of cerebral palsy | GMFCS* | MoCA | SF36/ average rating | Physical activity level | Education | Employment status (student, working) |
| --- | --- | --- | --- | --- | --- | --- | --- | --- | --- |
| CP_01 | 21-25 | F | spastic right sided hemiparesis | level I | 26 | 83.1 | low | incomplete higher education | student |
| CP_02 | 21-25 | M | spastic right sided hemiparesis | level I | 28 | 88.4 | low | incomplete higher education | student |
| CP_03 | 21-25 | F | spastic diplegia | level II | 28 | 69.4 | low | incomplete higher education | student |
| CP_04 | 21-25 | F | spastic diplegia | level III | 26 | 84.6 | low | incomplete higher education | working |
| CP_05 | 21-25 | F | spastic left-sided hemiparesis | level II | 29 | 87.8 | low | incomplete higher education | student |
| CP_06 | 31-35 | F | spastic diplegia | level III | 26 | 60.1 | low | higher education | working |
| CP_07 | 31-35 | F | spastic diplegia and hyperkinetic syndrome | level III | 26 | 74.4 | average | higher education | working |
| CP_08 | 26-30 | F | another type of cerebral palsy | level II | 28 | 85.1 | average | higher education | working |

**Note:**
M – male
F – female
\*Gross Motor Function Classification System (GMFCS) [19; 20]
Level I: Can walk indoors and outdoors and climb stairs without using their hands for support; can run and jump; has decreased speed, balance, and coordination.
Level II: Can walk indoors and outdoors and climb stairs using a railing; experiences difficulty with uneven surfaces, inclines, or while in crowds; can minimally run or jump.
Level III: Walks with assistive mobility devices indoors and outdoors on level surfaces; may be able to climb stairs using a railing; may propel a manual wheelchair; may require assistance for long distances or uneven surfaces.

The group of subjects with cerebral palsy included eight subjects (one man, seven women), with a mean age of 26 ± 3.61 years. Inclusion criteria were age over 18 years and the ability to independently stand on a stabilometric platform for the required duration of the experiment (with breaks provided upon request).

The control group included eight healthy adults (three men, five women), with a mean age of 23 ± 1.65 years. Inclusion criteria were the absence of a history of neurological or psychiatric diseases at the time of the study, as well as normal vision, hearing, and vestibular function.

The screening procedure for healthy volunteers included the MoCA (Montreal Cognitive Assessment) questionnaire, tests for manual and pedal asymmetry, and a survey to assess daily physical activity. To assess baseline static stability, a Romberg test was performed with eyes closed in three positions with a sequential decrease in the support surface [18].

Participants with cerebral palsy were tested using the MoCA questionnaire, the SF-36 quality of life questionnaire, and a survey on daily physical activity. Motor asymmetry tests were conducted in this group, but we considered them supplementary, as motor dominance could be forced by the underlying disease.

The results of the screening tests did not serve as an exclusion criterion.

### 2.2. Study design

The stabilogram was recorded using a Stabilan-01-2 device (Russia). The stabilogram sampling rate was 50 Hz. During the study, subjects stood barefoot on a platform with their eyes open. Subjects assumed a «European stance»: toes pointed outward at a 30° angle, arms relaxed at their sides. Subjects gazed at a 19-inch monitor positioned approximately 70 cm in front of the subject. A schematic representation of the experimental setup, identical to that used in this study, is presented in our previous publication [21]. Subjects with cerebral palsy assumed a comfortable posture and were asked, if possible, to position their feet so that their position was as close to the «European stance» as possible.

The baseline task (Baseline/Target) for all participants consisted of maintaining posture using real-time visual feedback. Subjects were required to hold a marker displaying the current position of the center of pressure of their feet in the center of a target on a monitor. For the healthy group, this condition was considered the reference level of postural stability in an ecologically valid situation (with a visual target and feedback).

Then, in addition to the baseline task, various types of cognitive load were sequentially presented. We used two contrasting types of cognitive load: verbal-logical (arithmetic and logical counting) and nonverbal, related to the processing of complex sound patterns (listening to the audio track «Noises of the big city», preferred music, or a fragment of Mozart’s «Eine kleine Nachtmusik» K. 525: I. Allegro).

Each experimental task lasted 60 seconds. The analysis strategy involved comparing stabilographic signal data during the Baseline/Target task with each task in which the Baseline/Target task was supplemented by cognitive load tasks in healthy subjects with similar performance to individuals with cerebral palsy.

### 2.3. Data Analysis

Center of pressure (COP) parameters measured using a force platform are traditionally used to quantify postural control. Standard time-domain metrics—such as statokinesiogram length and area or average COP velocity—reflect the overall efficiency of the sensorimotor system but do not reveal its underlying mechanisms.

In this study, power analysis of the COP in selected frequency bands was used to investigate the mechanisms of postural control. This method is suitable because frequency analysis is a standard tool for decomposing the stabilographic signal into physiologically interpretable components and allows us to move beyond the description of general instability to an analysis of the mechanisms underlying postural control. Data processing was performed using a specialized Python 3.8 script with the NumPy, Pandas, and SciPy libraries to implement classic power calculation algorithms in the specified frequency ranges. The data processing algorithm included the following steps: Data loading: Stabilographic data was loaded from text files containing the center of pressure (COP) coordinates along the X (frontal plane) and Y (sagittal plane) axes. Band-pass filtering: The original time signal was filtered using a set of 10 narrow-band Butterworth filters (range from 0.05 to 12.0 Hz).

The average signal power over the entire observation period (60 seconds) in each range was calculated using the formula:

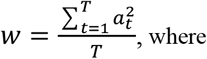

W is the signal power

t is the sample number (from 1 to T)

*α* is the signal amplitude

T is the total number of samples over the entire observation period.

Power calculations in the sagittal and frontal planes were performed separately.

The calculation of the stabilographic signal power was carried out in 10 frequency ranges from 0.05 to 12.0 Hz (F1: 0.05–0.1; F2: 0.1–0.5; F3: 0.5–1.0; F4: 1.0–1.5; F5: 1.5–2.0; F6: 2.0–4.0; F7: 4.0–6.0; F8: 6.0–8.0; F9: 8.0–10.0; F10: 10.0–12.0 Hz), taking into account the useful signal band of the stabiloplatform [22].

The range boundaries were defined in accordance with literature data linking specific frequency bands with activity at various levels of sensory control—from higher cortical functions (low frequencies, ∼0.05–0.5 Hz) and cerebellar-brainstem mechanisms (medium frequencies, ∼0.5–2.0 Hz) to spinal reflexes and biomechanical resonances (high frequencies, >2.0 Hz) [8–14]. Narrow ranges in the low-frequency region (e.g., 0.05–0.1 Hz) allow for a more detailed differentiation of the contribution of conscious control systems.

The structure of the ranges reflects the fact that the most clinically and physiologically significant information in the stabilogram is concentrated in the low-frequency part of the spectrum. Therefore, it is allocated a higher resolution (a greater number of narrow bands), while the wider high-frequency bands (above 4 Hz) correspond (according to the literature) mainly to noise and uninformative biomechanical vibrations. This approach is standard in frequency analysis of postural control.

### 2.4. Statistical Analysis of Power Indicators

To analyze the stabilographic signal power in the selected frequency ranges, two independent statistical approaches were used and compared to select the most adequate and informative method. Before comparing all indicators, the Shapiro-Wilk test (*α*=0.05) was used to check for normality of distribution. Depending on whether normality was observed, appropriate parametric or nonparametric statistical procedures were used for each of the two methods. The False Discovery Rate (FDR) method was used to control for errors in multiple comparisons.

Description of Comparison Methods:

Method 1 (Comparison with the Mean Value of the Healthy Group). This approach is based on assessing the position of the individual stabilogram power value in participants with cerebral palsy relative to the arithmetic mean of the healthy group. The «normal range» of values was defined in two ways: parametrically, as the mean ± 2 standard deviations (SD), which corresponds to an approximate 95% confidence interval assuming a normal distribution; and nonparametrically, as the interval between the 2.5 and 97.5 percentiles. Z-scores and p-values were used for quantitative assessment, and the Z-test (for a normal distribution) or its nonparametric rank analogs (e.g., the signed-sign test) were used for hypothesis testing.

Method 2 (Comparison with the Median of a Healthy Group). This method compared the individual stabilogram power values of participants with cerebral palsy with the median of a healthy group, ensuring robustness to distribution asymmetry and outliers. The «normative range» was determined nonparametrically by constructing a bootstrap confidence interval (95% CI) for the median. To assess the statistical significance of individual value deviations, an approach was developed comprising three components: an adapted Wilcoxon test for assessing the rank position (extremity) of a patient’s observation relative to the entire healthy sample; a binomial test for assessing the direction of the deviation (e.g., the probability of values above the median being predominant); and a bootstrap procedure for assessing the accuracy and robustness of the resulting median and confidence limits. The key metrics were the absolute deviation from the median and the p-value reflecting the results of these three tests. Results were visualized using a violin plot, which simultaneously displays the empirical density distribution of data in the healthy group and the position of the median.

Thus, the two methods were implemented in parallel analysis streams, which will allow for further comparison of their sensitivity, robustness to distributional distortions, and the meaningful interpretability of the obtained results.

## 3. Results and Discussion

The initial analysis, conducted within the framework of the traditional model linking broad frequency ranges with the activity of individual sensory systems [8–14, 23–28], revealed not only the expected differences but also complex, difficult-to-interpret patterns of change. Marked shifts in power in narrow frequency bands could not be adequately explained by a simple strengthening or weakening of any single sensory modality. The observed pattern rather indicated that the changes were not local (modality-specific) but rather global and systemic in nature, affecting the organization of the motor act as a whole. Explaining these data required a shift to a concept describing the holistic architecture of movement control. N.A. Bernstein’s level theory of movement construction [15] was chosen as the theoretical basis, allowing for the interpretation of the frequency components of the stabilogram as a manifestation of the activity of various neuromotor control circuits—from spinal to cortical. This allowed us to develop a model in which each narrow frequency range is considered as a potential indicator of the functional contribution of a certain level of motor act organization. We associated the level of higher cortical functions (E-level according to Bernstein) with a frequency of 0.05–0.1 Hz; the parietal-premotor level (D-level according to Bernstein) with a frequency of 0.1– 0.5 Hz; the pyramidal-striatal level (C1 and C2 according to Bernstein) with 0.5–1.0 Hz and 1.0–1.5 Hz; the thalamo-palidal (B-level according to Bernstein) with 1.5–2.0 Hz; and the rubrospinal level (A-level according to Bernstein) with 2.0–4.0 Hz, 4.0–6.0 Hz, 6.0–8.0 Hz, 8.0–10.0 Hz, and 10.0– 12.0 Hz (Fig. 1).

**Figure 1.**
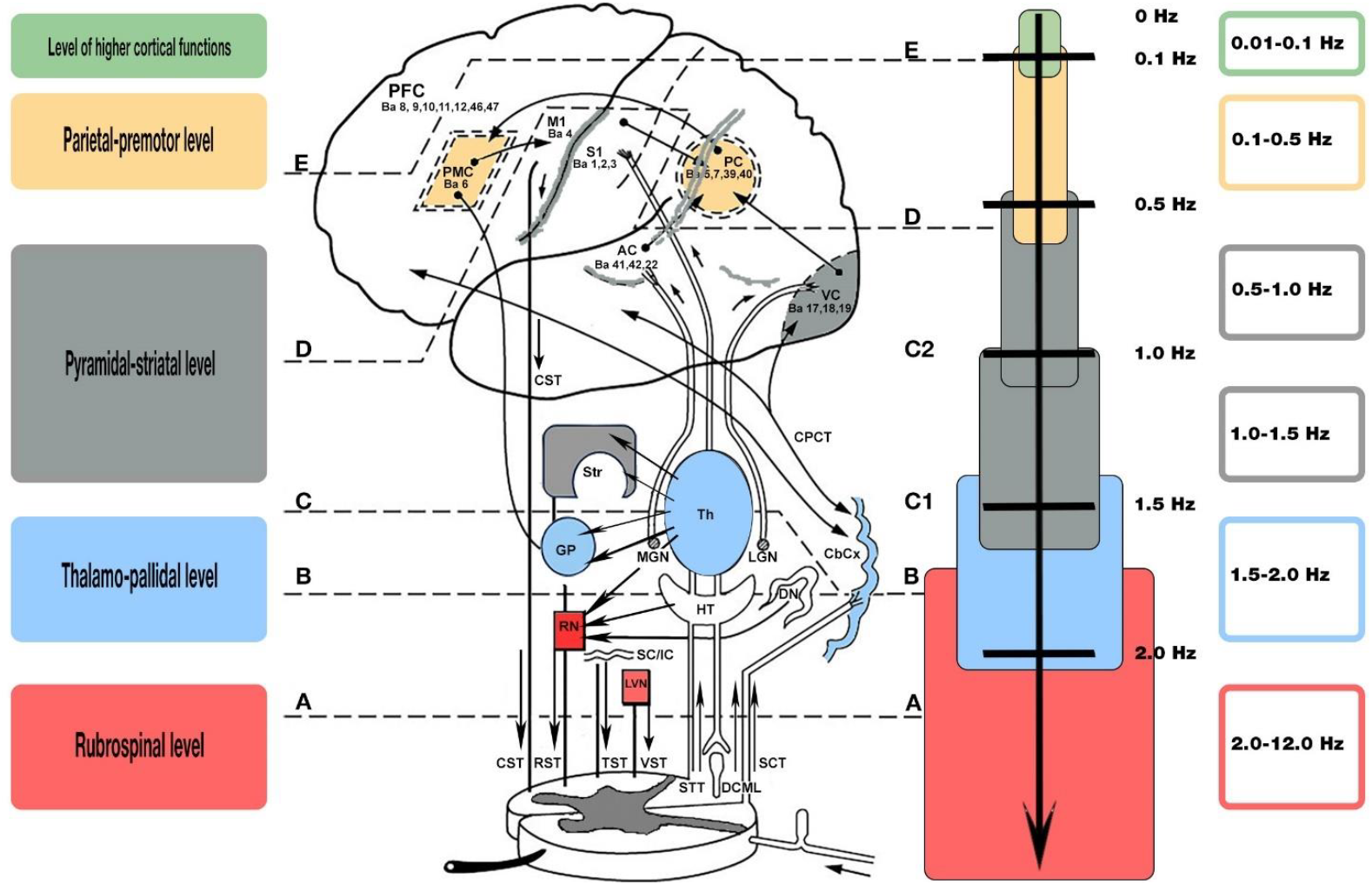
N.A. Bernstein’s model of levels of movement control with a schematic distribution of the conducting pathways and nuclei of the brain (modified and revised). The original diagram is presented in N.A. Bernstein’s book «On the Construction of Movements» Medgiz, 1947, p. 154, Fig. 79 [15]. Ba — Brodmann areas; CST — Corticospinal Tract; RST — Rubrospinal Tract; TST — Tectospinal Tract; VST — Vestibulospinal Tract; STT — Spinothalamic Tract; DCML — Dorsal Column-Medial Lemniscus pathway; SCT — Spinocerebellar tracts; LVN — Lateral Vestibular Nucleus; SC/IC — Superior Colliculi/Inferior Colliculi; RN — Red Nucleus; HT — Hypothalamus; DN — Dentate Nucleus; GP — Globus Pallidus; Th — Thalamus; MGN — Medial Geniculate Nucleus; LGN — Lateral Geniculate Nucleus; CbCx — Cerebellar Cortex; Str — Striatum; CPCT — Corticopontocerebellar pathway; VC — Visual Cortex, Brodmann areas 17, 18, 19; AC — Auditory Cortex, Brodmann areas 41, 42, 22; PC — Parietal Cortex, Brodmann areas 5, 7, 39, 40; S1 — Primary Somatosensory Cortex, Brodmann areas 1, 2, 3; M1 — Primary Motor Cortex, Brodmann areas 4; PMC — Premotor Cortex, Brodmann areas 6; PFC — Prefrontal cortex, Brodmann areas 8, 9, 10, 11, 12, 46, 47

Changes in the strength of the stabilometric signal within the framework of our proposed model allowed us to construct individual “impairment maps” of postural control in individuals with cerebral palsy. In Table 3, statistically significant deviations are highlighted in red; these levels can be interpreted as zones of functional overload or deficit in the motor control hierarchy. Postural control in the frontal and sagittal planes is regulated by different muscle synergies and is largely independent [29]. According to current concepts, the central nervous system establishes muscle tone at key balance control points to ensure constant and sufficient stiffness in the frontal plane to counteract inertial and gravitational forces [30]. Due to this specificity, oscillations in the frontal plane can reveal more complex and hidden pathological patterns that require their separate analysis.

**Table 3.** Comparison of the stabilogram power values of the healthy group (median) and the individual values of participants with CP (CP_001-CP_008) for oscillations in the sagittal and frontal planes during the task. 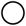 P-value>0.05,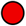P-value<0.05 (Method 2 was used—comparison with the median of the healthy group).

| Bernstein level | Freq., Hz | Level of oscil. | Health | CP_001 | CP_002 | CP_003 | CP_004 | CP_005 | CP_006 | CP_007 | CP_008 |
| --- | --- | --- | --- | --- | --- | --- | --- | --- | --- | --- | --- |
| Level of higher cortical functions | 0.05–0.1 | Front. | 0.2140 | ○ | ○ | ○ | ○ | ○ | ● | ● | ○ |
|  |  | Sagit. | 0.5119 | ○ | ○ | ○ | ○ | ○ | ○ | ○ | ○ |
| Parietal-premotor level | 0.1-0.5 | Front. | 1.4022 | ○ | ○ | ○ | ● | ○ | ○ | ● | ● |
|  |  | Sagit. | 1.9811 | ○ | ○ | ○ | ● | ○ | ○ | ● | ● |
| Pyramidal -striatal level | 0.5-1.0 | Front. | 0.6088 | ○ | ○ | ○ | ● | ○ | ○ | ● | ● |
|  |  | Sagit. | 0.5930 | ○ | ○ | ○ | ● | ● | ○ | ● | ● |
|  | 1.0-1.5 | Front. | 0.0913 | ○ | ○ | ○ | ● | ○ | ○ | ● | ● |
|  |  | Sagit. | 0.1057 | ○ | ○ | ○ | ● | ○ | ○ | ○ | ○ |
| Thalamo-pallidal level | 1.5-2.0 | Front. | 0.0144 | ○ | ○ | ○ | ● | ○ | ○ | ● | ○ |
|  |  | Sagit. | 0.0275 | ○ | ○ | ○ | ● | ○ | ○ | ● | ● |
|  | 2.0-4.0 | Front. | 0.0136 | ○ | ○ | ○ | ● | ○ | ○ | ● | ● |
| Rubrospinal level |  | Sagit. | 0.0274 | ● | ○ | ○ | ● | ○ | ○ | ● | ● |
|  | 4.0-6.0 | Front. | 0.0011 | ○ | ○ | ○ | ● | ○ | ○ | ○ | ● |
|  |  | Sagit. | 0.0038 | ● | ○ | ○ | ● | ○ | ○ | ● | ○ |
|  | 6.0-8.0 | Front. | 0.0011 | ○ | ○ | ○ | ○ | ○ | ○ | ○ | ○ |
|  |  | Sagit. | 0.0030 | ○ | ○ | ○ | ○ | ○ | ○ | ○ | ○ |
|  | 8.0-10.0 | Front. | 0.0007 | ○ | ○ | ○ | ○ | ○ | ○ | ○ | ○ |
|  |  | Sagit. | 0.0029 | ○ | ○ | ○ | ○ | ○ | ○ | ○ | ○ |
|  | 10.0-12.0 | Front. | 0.0002 | ○ | ○ | ○ | ○ | ○ | ○ | ○ | ○ |
|  |  | Sagit. | 0.0004 | ○ | ○ | ○ | ○ | ○ | ○ | ○ | ○ |

**Note:**
Frequency — Freq.; Level of oscillations — Level of oscil.; Sagittal — Sagit.; Frontal — Front.

To illustrate in detail the disturbances summarized in the postural control maps, we present an example of the analysis of the stabilogram power in the frequency range of 0.05–0.1 Hz (Figure 2), which, in our hypothesis, is associated with the activity of higher cortical functions. This range was chosen for demonstration as the most indicative in the context of compensatory mechanisms in cerebral palsy [31; 32]. The figure presents data for a group of healthy participants and participants with cerebral palsy, obtained while performing the Baseline/Target task in the sagittal and frontal planes. The analysis revealed a statistically significant (p<0.05) increase in the oscillation power in the sagittal plane in this range in two participants with cerebral palsy (CP_006 and CP_7). We interpret this observation as a sign of pathological hyperactivity of the level of conscious control (E-level according to Bernstein). To maintain an upright posture, the brain of an adult with cerebral palsy is forced to intensively engage higher cortical functions—attention, volitional effort, and constant conscious control. The automated, subcortical mechanisms of balance maintenance (levels D, C, and B), which operate in the background in a healthy individual, are damaged or functionally insufficient in cerebral palsy. To compensate for this deficit, the cerebral cortex takes on the unusual, routine task of postural stabilization, functioning in a chronically overstrained state. Thus, maintaining an upright posture for an adult with cerebral palsy is transformed from an automatic process into an active, extremely resource-intensive task requiring continuous conscious control [31–34]. This directly explains the high «cost» of maintaining balance in cerebral palsy, clinically manifested in rapid fatigue and significant difficulties in performing parallel tasks (e.g., speaking or manipulating objects)—cognitive resources are primarily «diverted» to postural control. The presented frequency range is especially important because the low power of the stabilogram in it in other participants with cerebral palsy indicates a different pathological pattern, in which the task is not transferred to the level of conscious control but remains solvable at lower, automated, and probably pathologically altered levels.

**Figure 2.**
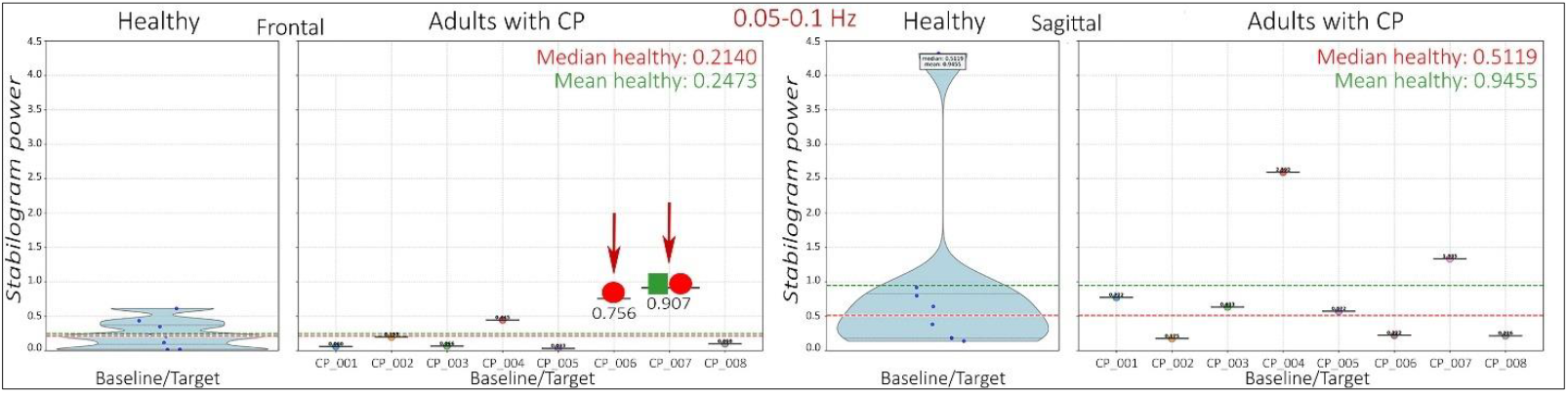
Power distribution in the frequency range 0.05-0.1 Hz (Bernstein E-level) in healthy participants and participants with cerebral palsy during oscillations in the frontal and sagittal planes during the Baseline/Target task. 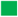P-value < 0.05 (Method 1 was used - comparison with the average power value of the healthy group). 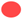P-value < 0.05 (Method 2 was used - comparison with the median power of the healthy group).

Thus, the individual frequency profiles of participants with cerebral palsy in the Baseline/Target task revealed a pattern of overcompensation, manifested by an increase in stabilogram power in the 0.05-0.1 Hz range, which we associate with the level of higher cortical levels.

Because the Baseline/Target task is highly ecological, integrating vision, external reference points, and active motor control, it serves as a control potential for the system, demonstrating how a person utilizes all available resources to maintain posture. The next logical step in our analysis was to study how the postural control system responds to the additional task, that is, how the individual frequency profile changes with the addition of cognitive load. By adding different types of load, we can observe which components of the individual postural control profile (i.e., which levels of regulation) are most vulnerable or, conversely, are compensatorily activated.

Next, we present three representative cases illustrating specific changes in the individual profiles of participants with CP during cognitive tasks.

Figure 3 shows a statistically significant increase in stabilogram power in the 0.1-0.5 Hz frequency range—a range that, according to our hypothesis, corresponds to parietal-premotor activity (Bernstein’s level D). This increase was observed when comparing the performance of the basic postural maintenance task (Baseline/Target) with a condition in which postural maintenance was combined with arithmetic calculation. We interpret this pattern as a manifestation of compensatory hyperactivation: to simultaneously solve a postural and cognitive task, the system is forced to further intensify the work of higher cortical coordination mechanisms. We have termed these changes a «compensatory hyperactivation pattern».

**Figure 3.**
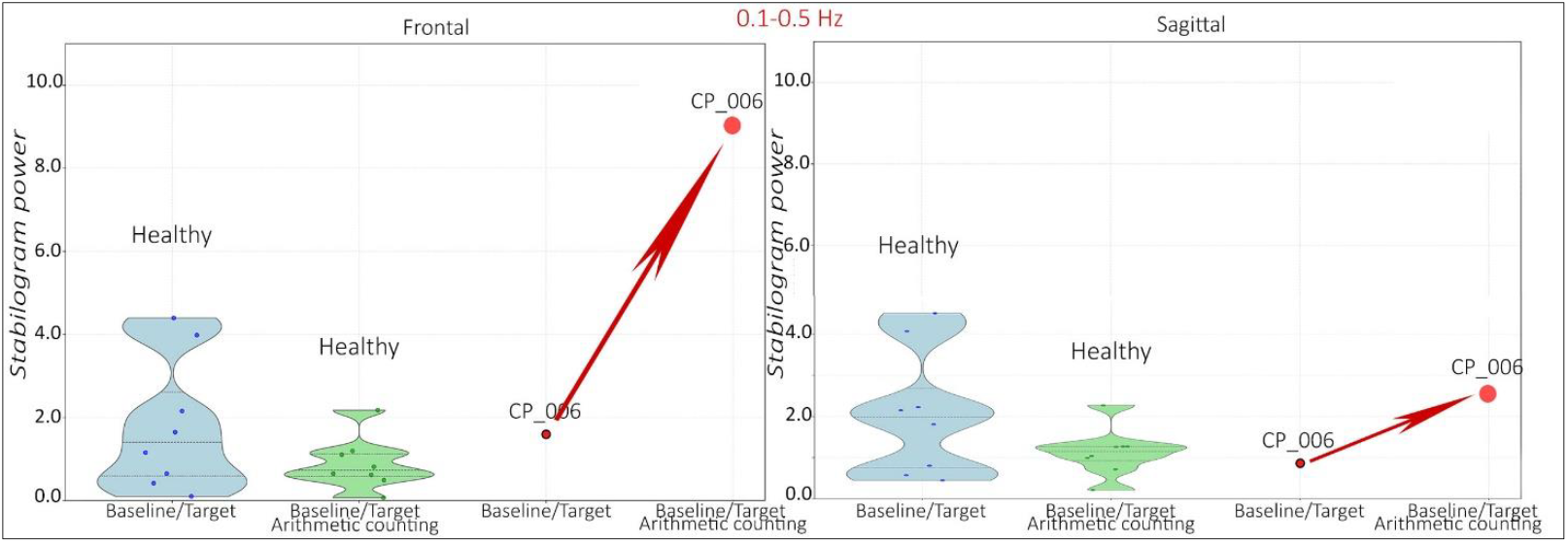
«Pattern of compensatory hyperactivation». Power distribution in the frequency range 0.1-0.5 Hz (Bernstein D level) in healthy participants and participants with cerebral palsy (CP_006) during oscillations in the frontal and sagittal planes during the performance of the Base/Target task and the Base/Target task supplemented by solving an arithmetic problem. 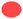P value <0.05 (method 2 was used - comparison with the median power of the healthy group).

Figure 4 shows a trend toward decreased power in the 1.0-1.5 Hz range, which corresponds to the pyramidal-striatal level (C2 according to Bernstein), when comparing the performance of the baseline posture maintenance task (Baseline/Target) with the condition in which the baseline task was accompanied by listening to Mozart’s music. Visually, this pattern is consistent with the hypothesis of «functional depletion» of resources at this level. However, the current sample size does not allow us to establish the statistical significance of this particular observation (P-value > 0.05). We have termed these changes the «functional depletion pattern». We believe that the appearance of this pattern serves as a landmark, clearly demonstrating functional depletion. However, to obtain reliable criteria for this pattern, a larger sample is necessary. This finding emphasizes the need for a larger sample for its validation.

**Figure 4.**
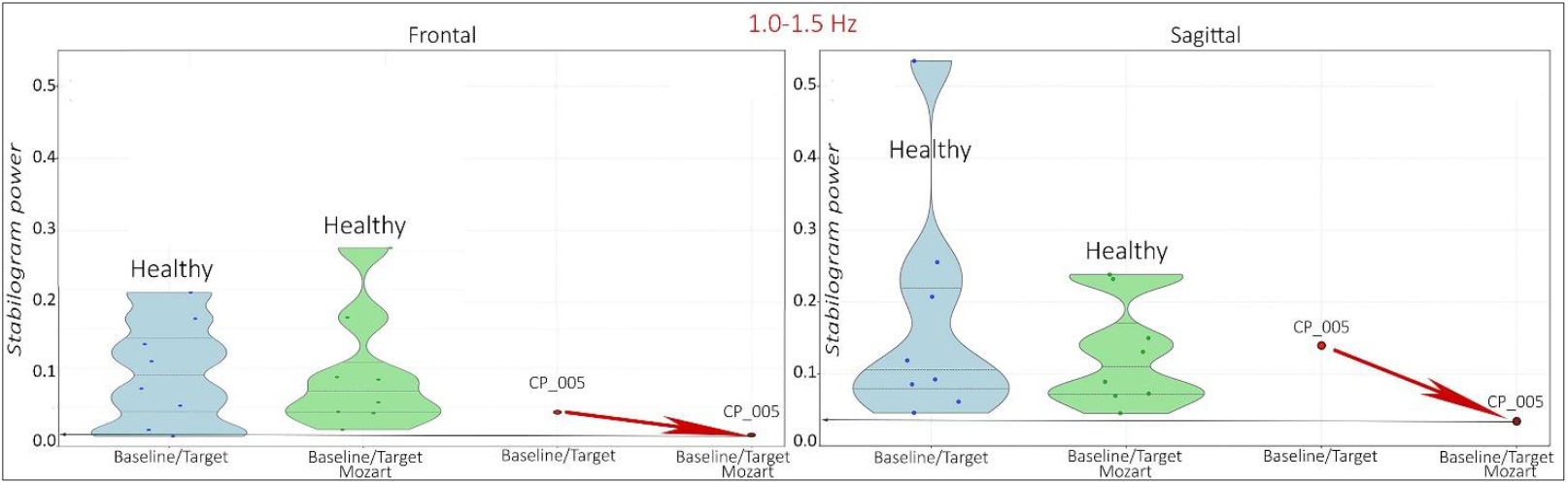
«Functional exhaustion pattern». Power distribution in the 1.0-1.5 Hz frequency range (Bernstein level C2) in healthy participants and in participants with cerebral palsy (CP_005) during oscillations in the sagittal and frontal planes while performing the Baseline/Target task and the Baseline/Target task supplemented with listening to Mozart’s music. P-value>0.05 (Method 2 was used — comparison with the median power of the healthy group).

Figure 5 shows individual changes in the stabilometric signal power toward normalization. When listening to Mozart’s music, a decrease in the stabilometric signal power in the 2.0-4.0 Hz range, which in our hypothesis corresponds to the rubrospinal level (Bernstein level A), is observed, approaching normal values. This effect suggests that the structured musical stimulus in this case acts not as a distracting load but as an external «pacemaker» modulating the activity of the most basic, spinal level of regulation, reducing pathological muscle tension and excessive reflex oscillations. We have termed these changes the «normalization pattern».

**Figure 5.**
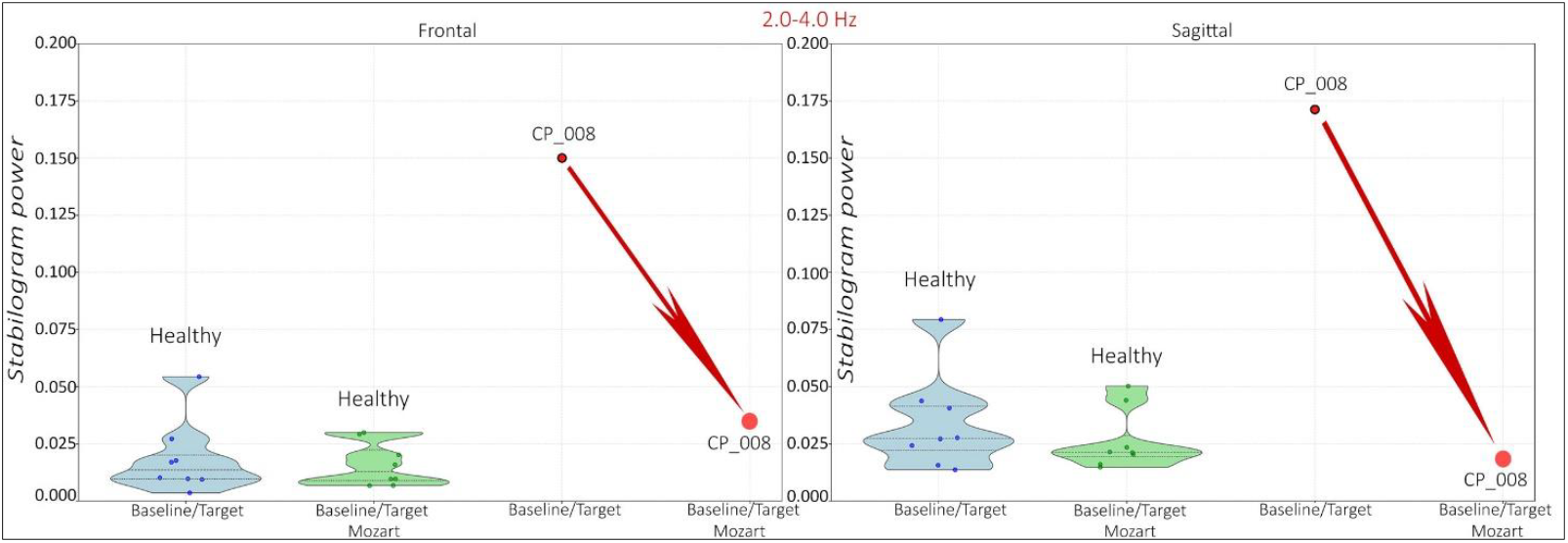
«Normalization Pattern». Distribution of power in the frequency range 2.0-4.0 Hz (Bernstein level A) in healthy participants and in participants with cerebral palsy (CP_008) during oscillations in the frontal and sagittal planes during the Baseline/Target task and the Baseline/Target task supplemented with listening to Mozart music. 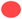P-value < 0.05 (Method 2 was used — comparison with the median power of the healthy group).

Thus, the presented data demonstrate possible changes in the stabilometric signal strength, reflecting the postural control system’s responses to cognitive load in cerebral palsy. These changes lead to compensatory hyperactivation of higher levels of regulation, functional exhaustion of intermediate components of postural control, and optimization of basic levels under the influence of an external acoustic stimulus (patterns of «hyperactivation», «exhaustion», and «optimization»). Interpretation of the results within the framework of N.A. Bernstein’s level theory of movement construction allows us to consider these patterns as a manifestation of imbalance or synergy between various postural regulation circuits—from subspinal to corticocerebellar. Of particular significance is the identified optimization response, indicating that the system’s preserved neuroplastic potential can be specifically engaged to normalize postural control through specific modulating effects.

## Limitations of the study

The main limitations of the study include the small sample size and its non-clinical nature (the lack of detailed clinical characteristics of the participants by type and severity of cerebral palsy). Furthermore, a single patient may simultaneously exhibit multidirectional changes at several levels of postural control, leading to the formation of a complex individual profile combining signs of hyperactivation and deficit. Although this directly points to the need for personalized rehabilitation approaches, the data from this pilot study do not allow for the identification of established physiological clusters or the formulation of rehabilitation recommendations. The obtained results primarily demonstrate the fundamental feasibility and diagnostic value of the approach, setting the direction for further research on a larger sample.

## 4. Conclusion

Developed based on N.A. Bernstein’s level theory [15], the frequency analysis method for stabilogram power allowed us to identify three key patterns of systemic reorganization of postural control in cerebral palsy: compensatory hyperactivation, functional exhaustion, and optimization of basic levels. This triad indicates that localized damage to the central nervous system leads to a global reorganization of the entire hierarchical movement control system with the formation of new compensatory functional connections [16; 17].

This study confirms that adults with cerebral palsy (CP) represent a pathophysiologically heterogeneous group, in which different localizations and severities of CNS damage create diverse impairment profiles within the level model of movement regulation. The data obtained indicate that these impairments are not universal, reflecting the wide clinical diversity of CP and emphasizing the need to identify clusters based on objective physiological markers. We hypothesize the existence of specific pathophysiological phenotypes within this group, manifested as unique individual postural control profiles. A characteristic feature of the identified phenotypes, regardless of the clinical type of cerebral palsy, will be the presence of three basic patterns of change: «hyperactivation», «exhaustion», and «optimization».

We believe that the identified patterns and changes in the stabilometric signal strength may form the basis for developing «personalized» rehabilitation approaches aimed not at general balance training, but at the targeted transformation of a specific pathological profile through the individual selection of modulating interventions (cognitive, sensory, and motor). It can be assumed that this approach also has significant rehabilitation potential for use in other CNS pathologies involving damage to the pyramidal tracts and subcortical structures (e.g., the consequences of traumatic brain injury).

## Data Availability

All data produced in the present study are available upon reasonable request to the authors

## Ethical Standards

All participants were informed in advance about the study’s aims and methods and provided informed consent. The study was conducted in accordance with the principles of the Declaration of Helsinki (1964) and its subsequent updates. The study protocols were approved by the local ethics committee of the Institute of Higher Nervous Activity and Neurophysiology of the Russian Academy of Sciences (Moscow) (Protocol No. 3 dated November 23, 2023, and Protocol No. 1 dated February 3, 2025).

## Authors’ contributions

Strelnikova E.V.: study conduct, formal analysis, development of the study hypothesis, writing, reviewing, and editing the manuscript.

Oknina L.B.: scientific supervision of the study, study conduct, development of the study hypothesis, writing, reviewing, and editing the manuscript.

Slezkin A.A.: formal analysis, development of the study hypothesis, writing, reviewing, and editing the manuscript.

Kantserova A.O.: writing, reviewing, and editing the manuscript. Myachina M.E.: study conduct.

Kirichenko A.V.: clinical and neurological examination of participants with cerebral palsy, interpretation of study results.

Shtern M.V.: clinical and neurological examination of participants with cerebral palsy, interpretation of study results.

